# Diagnostic testing in people with primary ciliary dyskinesia: an international participatory study

**DOI:** 10.1101/2023.01.03.23284153

**Authors:** Leonie D Schreck, Eva SL Pedersen, Isabelle Cizeau, Loretta Müller, Catherine Kruljac, Jane S Lucas, Myrofora Goutaki, COVID-PCD patient advisory group, Claudia E Kuehni

## Abstract

Diagnostic tests are important in primary ciliary dyskinesia (PCD), a rare disease, to confirm the diagnosis and characterize the disease. We compared diagnostic tests for PCD between countries worldwide, assessed whether people with PCD recall their tests, and identified factors associated with the use of tests. We used cross-sectional data from COVID-PCD—an international participatory cohort study collecting information directly from people with PCD. The baseline questionnaire inquired about tests used for PCD diagnosis. Using logistic regression, we investigated factors associated with measurement of nasal nitric oxide (nNO), biopsy for electron or video microscopy, and genetic testing. We included data from 747 participants (60% females) from 49 countries worldwide with median age 27 (interquartile range 12–44). Most (92%) reported diagnostic tests for PCD. Participants reported measurements of nNO (342; 49%), biopsy samples (561; 75%), and genetic tests (435; 58%). The reported use of individual tests, such as genetics, varied between countries from 38% in Switzerland to 68% in North America. Participant recall of test type also differed between countries with lowest recall in Switzerland. One-third (232; 36%) of participants reported all three tests (nNO, biopsy, and genetics). Recently diagnosed people reported more tests [nNO odds ratio (OR) 2.2, 95% Confidence Interval (CI) 1.5–3.2; biopsy OR 3.2, 95%CI 2.1–4.9; genetics OR 4.7, 95%CI 3.2–6.9] and those with situs abnormalities fewer tests (nNO OR 0.5, 95%CI 0.4–0.7; biopsy OR 0.5, 95%CI 0.4–0.8; genetics OR 0.7, 95%CI 0.5–0.94). Our results indicate PCD diagnostic testing differed widely around the world and many patients received incomplete diagnostic work-up based only on clinical features or single tests. People diagnosed long ago and those with situs abnormalities possibly benefit from supplementary testing to refine their diagnosis as a prerequisite for personalized medicine.

## Introduction

For people with primary ciliary dyskinesia (PCD), diagnostic tests are important to confirm the diagnosis and characterize the disease (1-3). PCD is a rare heterogeneous genetic disease with an estimated prevalence of around 1:7,500 to 1:10,000 (4, 5). Diagnosis relies on different tests, including measurement of nasal nitric oxide (nNO), assessment of ciliary function by high-speed video microscopy analysis (HSVA), visualisation of the ultrastructure by electron microscopy (EM) and immunofluorescence (IF), and genetic testing (6-8). Multiple tests are usually needed to diagnose PCD; however, for a proportion of individuals, genetic testing or EM are sufficient as single tests to confirm PCD (9, 10). Yet even among these people, a combination of different tests is important to understand associations between symptoms, defects in ciliary ultrastructure and function, and genotypes (1, 11). Different disease phenotypes have different prognoses, they require adaptations in treatment plans and monitoring (6, 12). Diagnostic characterization becomes increasingly important with regard to the development of personalized treatments (13) since only diagnostically well-characterized people with PCD will be eligible to participate in clinical trials and qualify for resulting treatments.

We know little about diagnostic testing for this rare disease in different countries. A survey among physicians from 194 paediatric PCD centres in 2007 looked at the availability of tests for PCD and showed that larger centres and those situated in countries with higher general government expenditures on health offered more tests (4, 14). An international patient survey performed in 2014 (15) and a study from the international PCD (iPCD) cohort that analysed data collected until 2018 (16) also evaluated diagnostic testing among patients with PCD. Both studies focused on patients with PCD in Europe. The iPCD study investigated a previously recommended test combination, but not genetic tests use (16). Diagnostic options for PCD have improved considerably over the past decade. Yet, we lack recent studies to understand what tests are actually performed, whether people with PCD know and remember tests, and what characteristics are associated with more comprehensive testing.

We analysed data from an ongoing international study of people with PCD to understand which diagnostic tests and test combinations were performed in various parts of the world, how well people with PCD recall their tests, and what factors explain using different tests.

## Materials and methods

### Study design and ethics

We used cross-sectional data from COVID-PCD—an ongoing participatory cohort study collecting information directly from people with PCD (clinicaltrials.gov registration number: NCT04602481). In collaboration with PCD patient support groups worldwide, the study was set up in spring 2020 to follow people with PCD throughout the COVID-19 pandemic and study other PCD-related research questions (17). COVID-PCD questionnaires are available in five languages (English, French, German, Italian, Spanish) and completed anonymously online. In COVID-PCD, people with PCD or their caretakers, such as parents of children with PCD, actively contribute to study design and questionnaire content. Patient support groups advertised the study and motivated members to participate. We extracted data for analysis on October 20, 2022.

The Bern Cantonal Ethics Committee (Kantonale Ethikkomission Bern) in Switzerland (study ID: 2020-00830) approved the study. Participants provided informed consent when they registered for the study. We report according to Strengthening the Reporting of Observational Studies in Epidemiology (STROBE) recommendations (18).

### Study procedures

When participants register for the study, they complete a baseline questionnaire asking about country of residence, symptoms and clinical problems, such as situs abnormalities, and details about their PCD work-up, such as diagnostic tests, age, and year of PCD diagnosis. Participants enter data directly into a web-based database using the Research Electronic Data Capture (REDCap) platform, hosted by the Swiss medical registries and data linkage centre (SwissRDL) at the University of Bern. We included all COVID-PCD participants who completed the baseline questionnaire in our study and reported to have PCD.

### Information on diagnostic testing and participant characteristics

Baseline questionnaires asked participants about diagnostic testing and use of specific tests (nNO, biopsy, and genetic testing). If participants reported biopsy, we asked whether their sample was analysed via high-speed video microscopy (HSVA), electron microscopy (EM), or both. All questions were accompanied by short explanatory texts describing test procedures. We classified missing and “I don’t know” responses as “no recall”.

Participants also reported characteristics in the baseline questionnaire. We grouped the countries of residence Scotland, England, Wales, and Northern Ireland as the United Kingdom (UK). We combined the United States and Canada as North America since they use the same diagnostic algorithm and their PCD care is organized in a network (10). We grouped countries with fewer than 25 participants together into either “other European countries” or “other non-European countries” (S1 Table). We categorized year of diagnosis into three periods (<=2000, 2001–2010, >=2011). We chose categories because of scarcity of diagnostic work-up before 2000, increased organization of PCD centres after 2010 (19), and the published consensus statement of the European Respiratory Society Task Force on PCD in children in 2009 (20). We asked participants about organ situs. If they reported any organs in different positions, we classified this as situs abnormalities. We labelled missing values as “not reported”. We provide the questions in S2 Table.

### Statistical analyses

We described use and recall of diagnostic tests by country and region. For nNO, we only included participants aged >=5 years—the age when this test is recommended and reliable (12). We studied factors associated with reported tests (nNO, biopsy, and genetic testing) using multivariable logistic regression. Outcome was coded as 1 if participants reported that tests were performed; if participants answered “no”, “I don’t know”, or it was not reported, we coded as 0. We included age at diagnosis, year of diagnosis, situs abnormalities, and country of residence as explanatory variables. For situs abnormalities, we combined the categories “no”, “I don’t know”, and “not reported” as “normal situs”. We performed sensitivity analyses assuming the “no recall” group had the test done. To do so, we combined the “no recall” group with the group that indicated that the test was performed and compared them with people who reported no test. We used R version 4.2.0 for all analyses.

## Results

### Characteristics of the study population

We included 747 people of whom 446 (60%) were female. The median age at survey was 27 years [interquartile range (IQR) 12–44, range 0–85 years] (Table 1). Study participants came from 49 countries, most commonly from North America (158; 21%), the UK (150; 20%), and Germany (107; 14%). The median age at diagnosis was 8 years (IQR 2–19). About half (347; 46%) of participants were diagnosed after 2010, one-third (231; 31%) before 2001, 146 (20%) between 2001—2010, and 23 (3%) participants did not report year of diagnosis. About half (345; 46%) of participants reported situs abnormalities.

**Table 1.** Demographic information and clinical characteristics of the study population of people with primary ciliary dyskinesia (COVID-PCD study, N = 747)

|  |  | n (%) |
| --- | --- | --- |
| **Sex** | Male | 299 (40) |
|  | Female | 446 (60) |
|  | Other | 2 (0) |
| **Age at survey** | Median (IQR, range) | 27 (12-44, 0-85) |
|  | 0-19 y | 270 (36) |
|  | 20-39 y | 240 (32) |
|  | 40-59 y | 192 (26) |
|  | > 60 y | 45 (6) |
| **Countries or regions<sup>a</sup>** | North America | 158 (21) |
|  | United Kingdom | 150 (20) |
|  | Germany | 107 (14) |
|  | Italy | 53 (7) |
|  | Switzerland | 48 (6) |
|  | France | 44 (6) |
|  | Australia | 33 (4) |
|  | Other European countries | 115 (15) |
|  | Other non-European countries | 39 (5) |
| **Age at diagnosis** | Median (IQR) | 8 (2-19) |
|  | Not reported | 23 (3) |
| **Year of diagnosis** | Before 2001 | 231 (31) |
|  | 2001-2010 | 146 (20) |
|  | After 2010 | 347 (46) |
|  | Not reported | 23 (3) |
| **Situs abnormalities** | Yes | 345 (46) |
|  | No | 386 (52) |
|  | I don't know | 15 (2) |
|  | Not reported | 1 (0) |
| **Congenital heart defect** | Yes | 58 (8) |
|  | No | 662 (89) |
|  | I don't know | 23 (3) |
|  | Not reported | 4 (1) |

Abbreviations: IQR, interquartile range. y, years <sup>a</sup>Countries with N≥25 displayed in table, countries with N<25 were categorised into other European countries and other non-European countries.

### Reported diagnostic testing

Of the 747 participants, most (690; 92%) reported diagnostic testing for PCD (S3 Table). Nasal NO had been measured in 342 of 693 participants older than age 5 (49%). Participants reported biopsy samples (561; 75%) for electron or video microscopy, and genetic tests (435; 58%) (Fig 1). Among those reporting biopsy, 325 (58%) stated their sample was visualised with HSVA and 283 (50%) told it was further analysed using EM (S3 Table).

**Fig 1.**
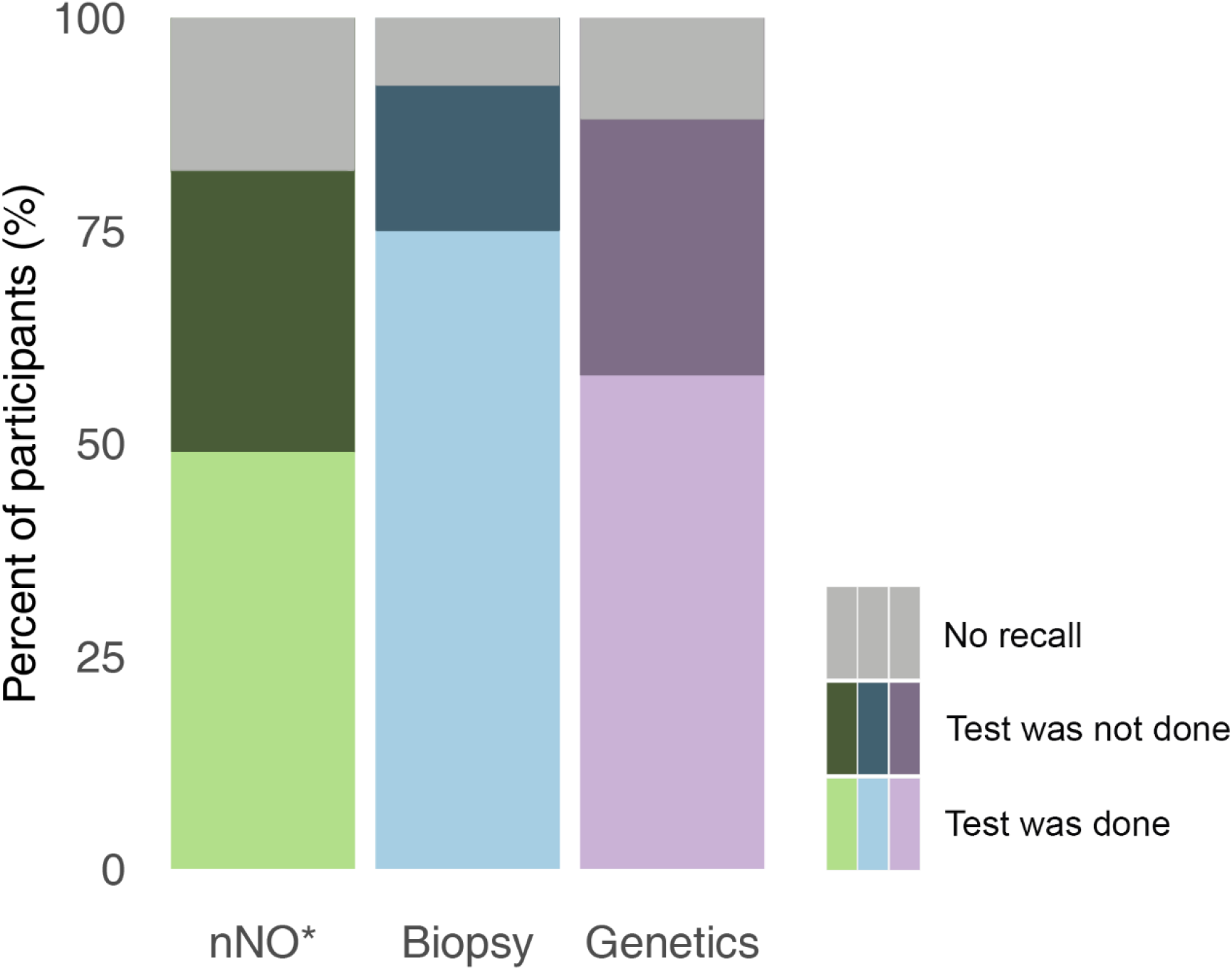
Diagnostic tests performed in people with primary ciliary dyskinesia (COVID-PCD study, N = 747). Abbreviations: nNO, nasal nitric oxide. *Only participants age >= 5 years are included (n = 693). Missing values and the answer category “I don’t know” were classified as “no recall” (missing values: nNO: n = 16, biopsy: n = 6, genetic testing: n = 9)

Reported tests differed between countries (Fig 2). Biopsy was the most common test in all countries except for North America and France where genetic testing dominated. For participants older than age 5, nNO measurements were reported by 66 (66%) of German participants, yet only by 15 (35%) people in Switzerland and 14 (36%) in other non-European countries. Biopsy samples were taken most often from people living in Australia (29; 88%), Italy (46; 87%), and the UK (128; 85%). Genetic testing was most often performed in France (31; 70%), Germany (73; 68%) and North America (108; 68%) and least often in Switzerland (18; 38%) and other non-European countries (17; 44%). Missing responses were fewer than 5% for all diagnostic tests.

**Fig 2.**
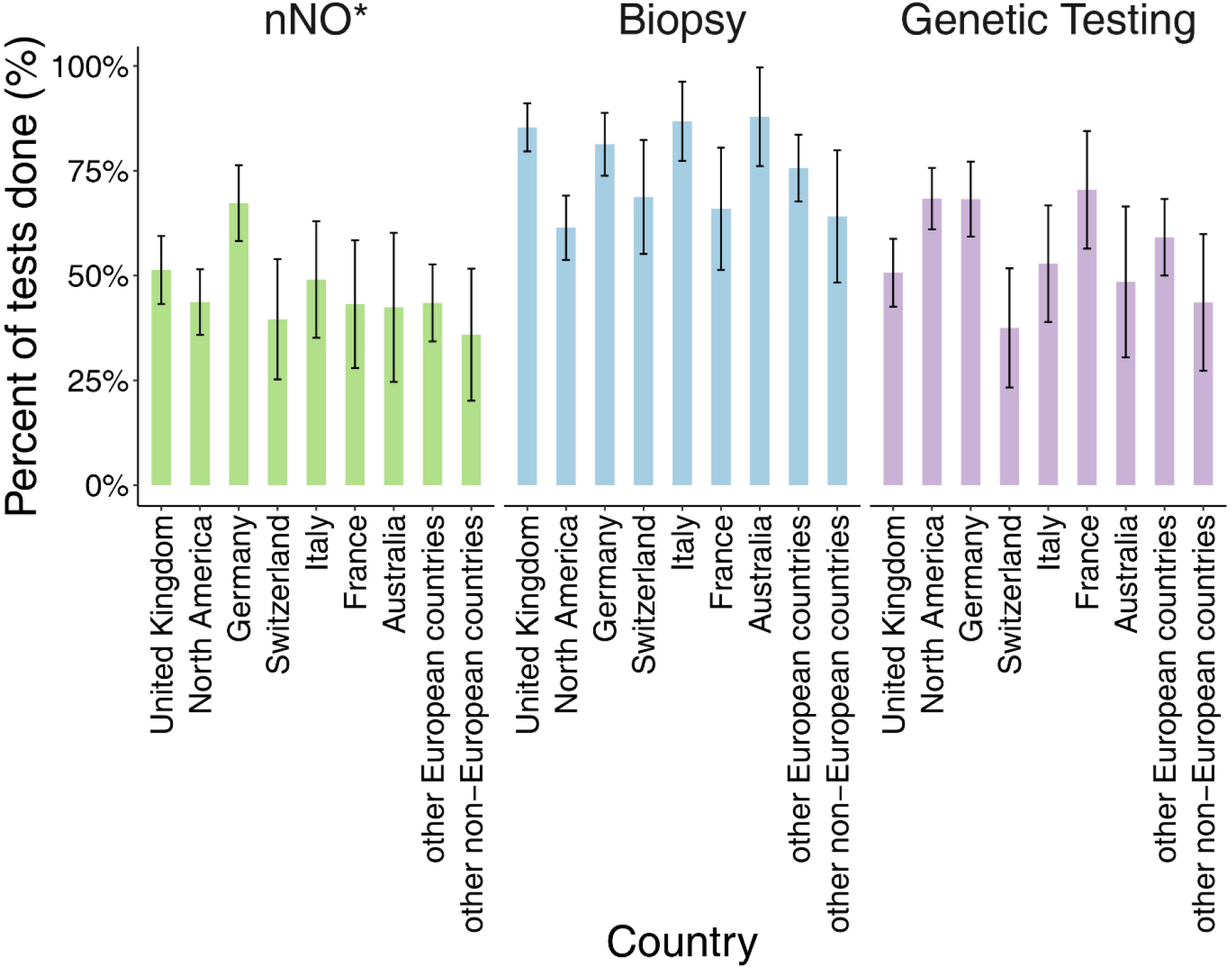
Diagnostic tests performed in people with primary ciliary dyskinesia, by country (COVID-PCD study, N = 747). Abbreviations: nNO, nasal nitric oxide. *Only participants age >= 5 years are included (n = 693).

### Recall of diagnostic testing

The proportion of people who did not remember whether a test was performed varied between tests (S3 Table); it was lowest for biopsy (57; 8%) and highest for nNO (125; 18%). Although people often knew that a biopsy sample was taken, they did not know whether it was analysed using HSVA (211; 38%) or EM (266; 47%).

The recall of tests also differed strongly between countries. Recall was poorest in Switzerland where 16 (37%) participants did not recall whether nNO was measured, 7 (15%) were unsure if biopsy samples were taken, and 10 (21%) did not know if genetic tests were performed.

### Combination of tests

Among participants older than age 5 with diagnostic tests, one-third (232; 36%) reported all three tests (nNO, biopsy, and genetics), 100 (16%) reported genetic testing and biopsy, and 101 (16%) reported biopsy alone (Fig 3). 57 (9%) did not remember which tests were performed. The frequency of combinations varied between countries. Most participants from Germany (53; 59%) reported combined analyses for nNO, biopsy, and genetic testing compared with only 7 (18%) in Switzerland and 6 (22%) in Australia (S3 Table). Of children younger than age 5, 39 (72%) reported the combination of biopsy and genetic testing.

**Fig 3.**
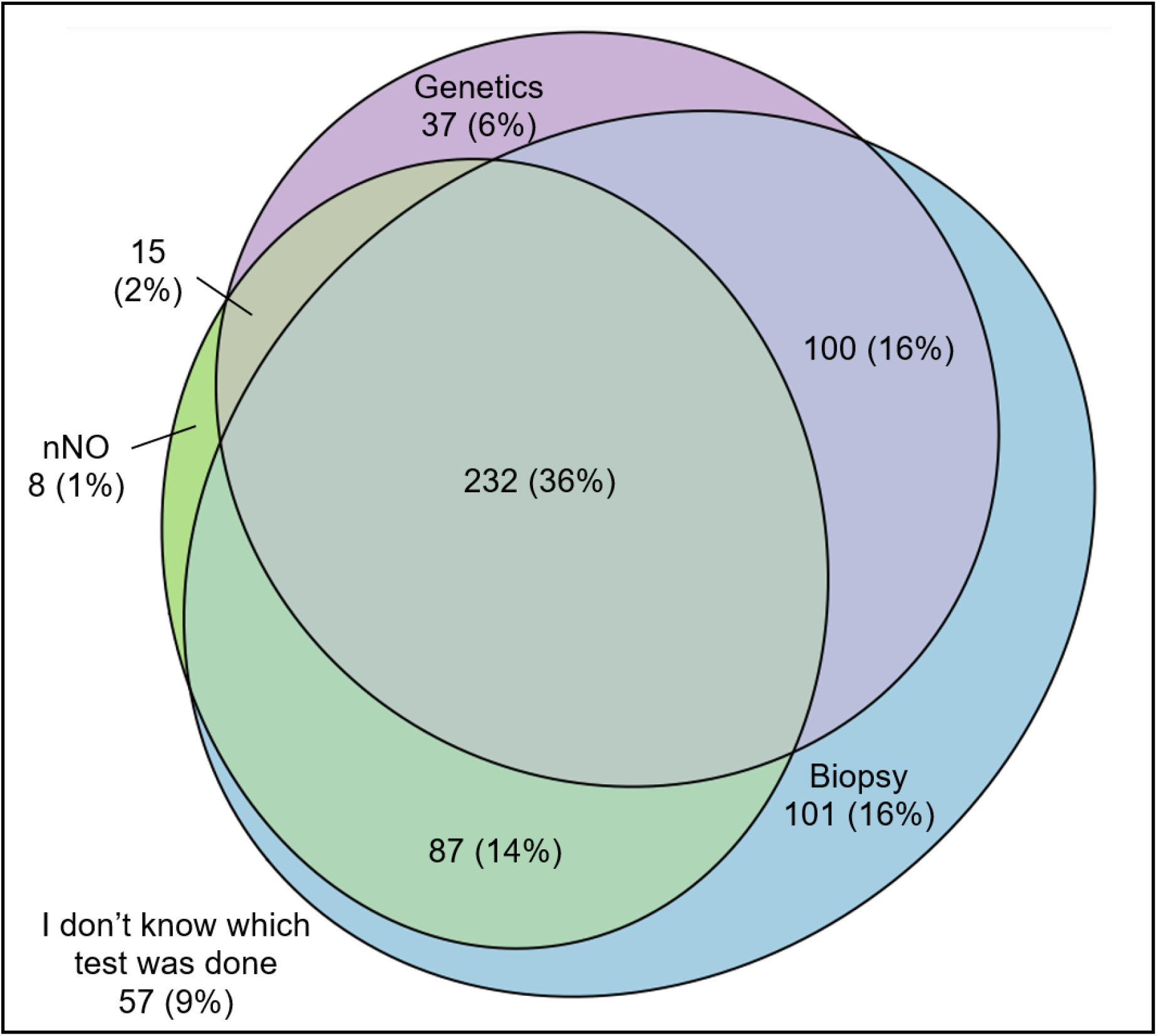
Venn diagram showing self-reported tests in people with primary ciliary dyskinesia who participated in COVID-PCD (*n* = 637) ^+^. Abbreviations: nNO, nasal nitric oxide. ^+^Participants under 5 years and/or who replied that no diagnostic was performed were excluded from this analysis.

### Predictors of diagnostic testing

Year of diagnosis and situs abnormalities were strongly associated with diagnostic testing (Fig 4). In a multivariable logistic regression, more people diagnosed after 2010 reported nNO measurement [Odds Ratio (OR) 2.2, 95% confidence interval (CI) 1.5–3.2], biopsy (OR 3.2, 95%CI 2.1–4.9), and genetic testing (OR 4.7, 95%CI 3.2–6.9) compared with people diagnosed before 2000. People with situs abnormalities reported fewer tests than people with situs solitus (nNO measurement: OR 0.5, 95%CI 0.4–0.7; biopsy: OR 0.5, 95%CI 0.4–0.8; genetic testing: OR 0.7, 95%CI 0.5–0.94). When compared with people from the UK, more Germans reported nNO measurements (OR 1.8, 95%CI 1.03–3.2), all countries (except for Italy) reported fewer biopsies, and participants from North America reported more genetic testing (OR 2.1, 95%CI 1.3–3.5). Age at diagnosis was not associated with any individual test; we excluded it from the final multivariable model (data not shown). Results from sensitivity analyses which assumed that everybody in the group “no recall” had the test done were similar to the main analysis (S5 Table).

**Fig 4.**
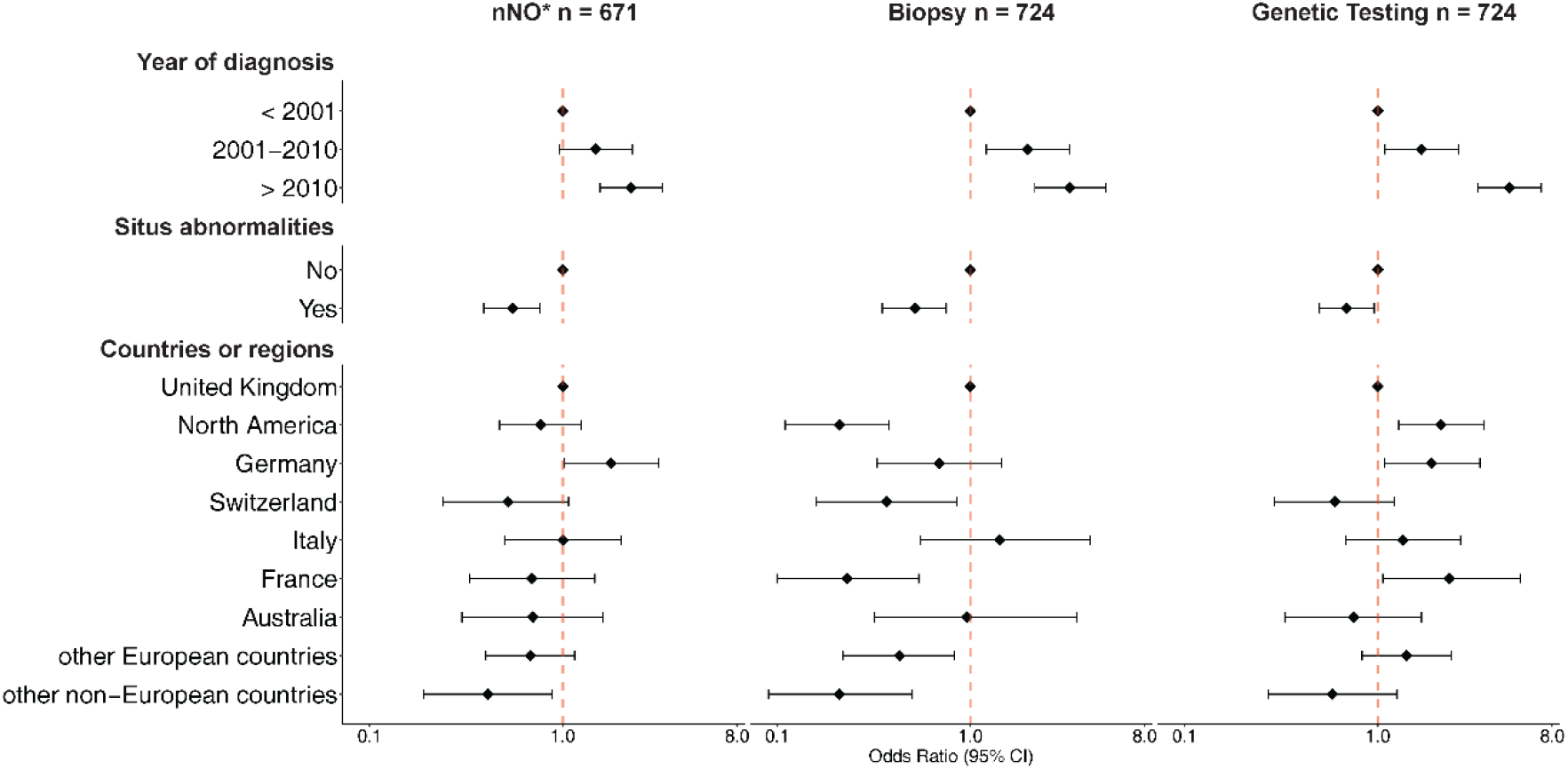
Factors associated with nNO measurement, biopsy and genetic tests in people with primary ciliary dyskinesia (COVID-PCD study). Abbreviations: CI, confidence interval. nNO, nasal nitric oxide. Participants who reported that the test was performed (“yes”) were compared to the group who reported either no test (“no”) or did not recall the test (“I don’t know” and missing). Odds Ratios were adjusted for all variables included in the model. *Only participants age >= 5 years are included.

## Discussion

### Summary of findings

Our international study of people with PCD found a large variation in diagnostic test use between countries. Even though most (92%) participants reported diagnostic testing for PCD, the use of individual tests varied widely. For example, only one-third of respondents from Switzerland reported genetic testing, whereas in North America two-thirds did. There were also differences in recall between countries. Overall, only one-third of participants older than age 5 reported all three tests (nNO measurement, biopsy, and genetics). People with PCD who were more recently diagnosed reported more tests, and those with situs abnormalities less.

### Strengths and limitations

With 747 enrolled participants, the COVID-PCD study is the largest study worldwide to collect data directly from people with PCD—it also enables comparisons between different regions of the world. The study includes the latest information about diagnostic testing among patients of all ages, including those who have not yet participated in clinical studies because they are treated by private physicians or reside in countries with decentralised PCD care. Another strength of the study is that it offers insight into what people with PCD know about their disease. However, self-reported data possibly leads to measurement bias, which is a limitation of our study. Since the study is anonymous, it was not possible to compare self-reported data with medical records. It might be participants were tested but not informed or forgot about tests. However, biopsies are uncomfortable, usually remembered procedures and genetic tests often need approval by patients and health insurance. When a nasal olive probe is inserted into one nostril during nNO measurement, it is also memorable. When we assumed tests were performed for all who reported no recall in a sensitivity analysis, the direction of the associations did not change, which strengthens the robustness of our findings. The COVID-PCD study invites all people with PCD to participate. Since the study was advertised by patient support groups, it may result in selection bias. Since patients involved with support groups are possibly better informed and treated than others who are not, our study may have underestimated the general lack of diagnostic work-up. However, the frequency of situs abnormalities and congenital heart defects (Table 1) in our study population was consistent with published data (21), suggesting participants were broadly representative in clinical terms.

### Comparison with other studies and interpretation

Genetic tests (58%) were more often reported in our study than in previous surveys. In a 2014 patient survey, only 39% reported genetic testing (15) and in the 2007 physician survey, 30% (4, 14). Due to the cost-effectiveness of multigene panels, genetic tests have recently become widely available. Also, the number of known disease-causing genes has steadily increased, so genetic testing can currently identify genes for more than 70% of people with PCD (12, 22). The large variation between countries reflects historical conditions or country-specific opportunities. In North America, genetic testing has been a cornerstone of the diagnostic algorithm for years, while in many European countries, PCD diagnosis has been based primarily on EM and HSVA. Projects such as the 100,000 Genomes Project in the UK led to a local increase of genetic tests (22-24). In other countries, availability of genetic testing is still limited for PCD diagnostics and rare diseases in general (25, 26). In many countries, structural changes in healthcare systems would be needed to improve access to genetic testing, which is a slow process that unfortunately limits care and equal opportunities for patients with rare diseases worldwide (27).

Recall of diagnostic tests varied between countries. It was overall lowest in Switzerland—the country with the second highest health expenditure per capita in the world since 1990, after the United States (28). PCD care in Switzerland is decentralised and many patients with PCD are treated by physicians who are not PCD specialists. Therefore, patients have insufficient information and education. For people with rare diseases, patient empowerment and shared decision-making are essential. For shared decision-making, patients must become experts as many encounter health care professionals with little expertise on their disease, particularly in emergency situations (29). Our findings indicate respondents’ knowledge about their disease is limited, which highlights the need for better information, education, and empowerment.

Although we showed test use differed between countries, diagnostic testing also depended strongly on year of diagnosis and situs. Age at diagnosis was not associated with the use of tests, which suggests diagnostic work-up is not more complete in paediatric compared with adult settings—a finding that contrasts better expected PCD awareness among paediatric pulmonologists. Our results suggest only newly diagnosed people with PCD benefitted from more comprehensive testing and newer tests, such as genetics. Although people diagnosed before 2000 often only received partial diagnostic work-up, they were not later recalled for supplementary testing to confirm and refine diagnoses. The same is true for people with PCD with situs abnormalities—physicians may be satisfied with clinical diagnosis and consider PCD proven. Thus, they do not offer biopsy or genetic testing to confirm diagnosis, which is insufficient because only 20–25% of people with situs inversus have PCD (30).

## Conclusion

We found PCD diagnostics differed markedly around the world and many people with PCD still have incomplete diagnostic work-up with diagnoses based only on clinical features or single tests. Several pre-clinical studies are developing novel molecular therapies for PCD; thus, understanding individual genotypes will become imperative for treatment. Therefore, it is important clinicians review tests and results with patients and plan further tests, if necessary. People diagnosed with PCD long ago and those with situs abnormalities possibly benefit from supplementary testing to improve diagnostic characterization as a prerequisite for personalized medicine.

## Data Availability

COVID-PCD data can be made available upon reasonable request by contacting Claudia Kuehni

## Acknowledgments

We thank all participants and their families, and we thank the PCD support groups and physicians who advertised the study. We thank our collaborators who helped set up the COVID-PCD study: Cristina Ardura, Yin Ting Lam, Christina Mallet, Helena Koppe, Dominique Rubi, University of Bern, and Amanda Harris, University Hospital Southampton. We thank the COVID-PCD advisory group (in alphabetical order): Sara Bellu, Associazione italiana Discinesia Ciliare Primaria Sindrome di Kartagener Onlus Italy; Isabelle Cizeau, Association ADCP, France; Fiona Copeland, PCD support UK; Katie Dexter, PCD support UK; Lucy Dixon, PCD support UK; Trini López Fernández, Asociación Española de Pacientes con Discinesia Ciliar Primaria, Spain; Susanne Grieder, Selbsthilfegruppe Primäre Ciliäre Dyskinesie, Switzerland; Catherine Kruljac, PCD Australia Primary Ciliary Dyskinesia, Australia; Michelle Manion, PCD Foundation, USA; Bernhard Rindlisbacher, Selbsthilfegruppe Primäre Ciliäre Dyskinesie, Switzerland; Hansruedi Silberschmidt, Verein Kartagener Syndrom und Primäre Ciliäre Dyskinesie, Germany. We thank the editorial service of the Institute of Social and Preventive Medicine at the University of Bern for the editorial suggestions.

## Ethics approval and consent to participate

The Bern Cantonal Ethics Committee (Kantonale Ethikkomission Bern) approved this study (Study ID: 2020-00830). Participants provide informed consent to participate in the study at registration into the study. Study participation is anonymous. Participants can withdraw their consent to participate at any time by contacting the study team.

## Availability of data and materials

COVID-PCD data can be made available upon reasonable request by contacting Claudia Kuehni

## Authors’ contributions

LD Schreck, ESL Pedersen, M Goutaki, and CE Kuehni made substantial contributions to the study concept and design and interpretation of the data. LD Schreck analysed the data and drafted the manuscript. LD Schreck, ESL Pedersen, I Cizeau, L Müller, C Kruljac, JS Lucas, M Goutaki, and CE Kuehni critically revised and approved the manuscript.

---

**S1 Table.** Countries of residence of COVID-PCD participants with less than 25 participants who were grouped into other European countries and other non-European countries.

| **Country** | **Number of participants** |
| --- | --- |
| **Other European countries** | **115** |
| Austria | 7 |
| Belgium | 9 |
| Croatia | 1 |
| Cyprus | 5 |
| Czech Republic | 1 |
| Denmark | 10 |
| Finland | 1 |
| Georgia | 2 |
| Greece | 1 |
| Hungary | 1 |
| Ireland | 9 |
| Jersey | 1 |
| Netherlands | 21 |
| Norway | 12 |
| Poland | 5 |
| Portugal | 3 |
| Romania | 1 |
| Spain | 16 |
| Sweden | 9 |
| **Other non-European countries** | **39** |
| Argentina | 1 |
| Bahrain | 2 |
| Brazil | 3 |
| Cameroon | 1 |
| Chile | 1 |
| Colombia | 1 |
| Ecuador | 1 |
| Hong Kong | 1 |
| India | 2 |
| Iran | 1 |
| Israel | 6 |
| Kuwait | 2 |
| Lebanon | 1 |
| Mexico | 2 |
| New Zealand | 1 |
| Panama | 1 |
| Puerto Rico | 2 |
| South Africa | 6 |
| Turkey | 3 |
| other country | 1 |

**S2 Table.** Formulation of questions and answers from the English adult baseline questionnaire of the COVID-PCD study.

| Question | Answer category |
| --- | --- |
| *Countries and regions* |  |
| Which country do you live in? | List of all countries worldwide |
| Which other country? | text |
| *Situs inversus* |  |
| Are any of your organs in a different position compared to most people? (E.g. the heart on the right side instead of on the left) | No<br>Yes<br>I don't know |
| *Diagnostic testing* |  |
| Have you had diagnostic tests for PCD? | No<br>Yes |
| Have you had a nasal nitric oxide test? (This test measures a gas from the nose through a thin tube that leads to a computer) | No<br>Yes<br>I don't know/I cannot remember |
| What was the nasal nitric oxide test result? | Normal<br>Suggestive of PCD (very low)<br>Borderline/unclear<br>I don't know/I cannot remember |
| Have you had a nasal brush biopsy? (Uncomfortable scraping or brushing to collect cilia/hair cells from the nose, or a brushing to collect cells from the airways during a bronchoscopy) | No<br>Yes<br>I don't know/I cannot remember |
| Do you know if the sample was tested by high speed video microscopy? (The sample was looked at under a microscope to see how the cilia/hairs move) | No, it was not tested by high speed video microscopy<br>Yes, it was tested by high speed video microscopy<br>I don't know/ I cannot remember |
| What was the result of the high speed video microscopy? | Static, slow, or abnormal movement, typical for PCD<br>Normal movement<br>Unclear result<br>I don't know/ I cannot remember |
| Do you know if the sample was tested by electron microscopy? (The sample was looked at with a microscope to see the inside structure of the cilia/hairs) | No, it was not tested by electron microscopy<br>Yes, it was tested by electron microscopy<br>I don't know/ I cannot remember |
| What was the result of the electron microscopy? | Typical for PCD<br>Normal<br>Unclear result<br>I don't know/I cannot remember |
| Have you had a genetic test (looking for genes that cause PCD)? | No<br>Yes<br>I don't know/ I cannot remember |
| Were any genes found that cause PCD? | No<br>Yes<br>I don't know/ I cannot remember/ waiting for results |
| *Year of diagnosis* |  |
| Which year were you diagnosed with PCD? | text (integer, Min: 1900, Max: 2025) |
| How old were you, when you were diagnosed with PCD? | text (integer, Min: 0, Max: 110) |

**S3 Table.** Diagnostic tests performed in people with primary ciliary dyskinesia (PCD), by country^a^ (COVID-PCD study, N = 747).

|  | Total<br>N = 747<br>n (%) | United<br>Kingdom<br>n = 150<br>n (%) | North<br>America<br>n = 158<br>n (%) | Germany<br>n = 107<br>n (%) | Italy<br>n = 53<br>n (%) | Switzerland<br>n = 48<br>n (%) | France<br>n = 44<br>n (%) | Australia<br>n = 33<br>n (%) | other<br>European<br>countries<sup>+</sup><br>n = 115<br>n (%) | other non-<br>European<br>countries<sup>+</sup><br>n = 39<br>n (%) |
| --- | --- | --- | --- | --- | --- | --- | --- | --- | --- | --- |
| **Any**<br>**diagnostic**<br>**test** |  |  |  |  |  |  |  |  |  |  |
| Done | 690 (92) | 144 (96) | 145 (92) | 97 (91) | 49 (92) | 43 (90) | 39 (89) | 32 (97) | 109 (95) | 32 (82) |
| Not done | 51 (7) | 4 (3) | 12 (8) | 8 (7) | 4 (8) | 4 (8) | 5 (11) | 1 (3) | 6 (5) | 7 (18) |
| No recall | 6 (1) | 2 (1) | 1 (1) | 2 (2) | 0 (0) | 1 (2) | 0 (0) | 0 (0) | 0 (0) | 0 (0) |
| **nNO<sup>b</sup>** | n = 693 | n = 145 | n = 142 | n = 100 | n = 49 | n = 43 | n = 41 | n = 28 | n = 106 | n = 39 |
| Done | 342 (49) | 76 (52) | 68 (48) | 66 (66) | 24 (49) | 15 (35) | 19 (46) | 13 (46) | 47 (44) | 14 (36) |
| Not done | 226 (33) | 39 (27) | 49 (35) | 22 (22) | 19 (39) | 12 (28) | 17 (41) | 11 (39) | 38 (36) | 19 (49) |
| No recall | 125 (18) | 30 (21) | 25 (18) | 12 (12) | 6 (12) | 16 (37) | 5 (12) | 4 (14) | 21 (20) | 6 (15) |
| **Biopsy** |  |  |  |  |  |  |  |  |  |  |
| Done | 561 (75) | 128 (85) | 97 (61) | 87 (81) | 46 (87) | 33 (69) | 29 (66) | 29 (88) | 87 (76) | 25 (64) |
| Not done | 129 (17) | 9 (6) | 47 (30) | 17 (16) | 6 (11) | 8 (17) | 9 (20) | 3 (9) | 19 (17) | 11 (28) |
| No recall | 57 (8) | 13 (9) | 14 (9) | 3 (3) | 1 (2) | 7 (15) | 6 (14) | 1 (3) | 9 (8) | 3 (8) |
| **Genetics** |  |  |  |  |  |  |  |  |  |  |
| Done | 435 (58) | 76 (51) | 108 (68) | 73 (68) | 28 (53) | 18 (38) | 31 (70) | 16 (48) | 68 (59) | 17 (44) |
| Not done | 223 (30) | 45 (30) | 37 (23) | 28 (26) | 16 (30) | 20 (42) | 11 (25) | 12 (36) | 34 (30) | 20 (51) |
| No recall | 89 (12) | 29 (19) | 13 (8) | 6 (6) | 9 (17) | 10 (21) | 2 (5) | 5 (15) | 13 (11) | 2 (5) |
| **HSVA<sup>c</sup>** | n = 561 | n = 128 | n = 97 | n = 87 | n = 46 | n = 33 | n = 29 | n = 29 | n = 87 | n = 25 |
| Done | 325 (58) | 61 (48) | 40 (41) | 73 (84) | 30 (65) | 22 (67) | 20 (69) | 21 (72) | 43 (49) | 15 (60) |
| Not done | 25 (4) | 3 (2) | 9 (9) | 2 (2) | 0 (0) | 0 (0) | 0 (0) | 1 (3) | 7 (8) | 3 (12) |
| No recall | 211 (38) | 64 (50) | 48 (49) | 12 (14) | 16 (35) | 11 (33) | 9 (31) | 7 (24) | 37 (43) | 7 (28) |
| **EM<sup>c</sup>** | n = 561 | n = 128 | n = 97 | n = 87 | n = 46 | n = 33 | n = 29 | n = 29 | n = 87 | n = 25 |
| Done | 283 (50) | 59 (46) | 46 (47) | 56 (64) | 26 (57) | 16 (48) | 14 (48) | 16 (55) | 38 (44) | 12 (48) |
| Not done | 12 (2) | 2 (2) | 1 (1) | 5 (6) | 0 (0) | 0 (0) | 0 (0) | 0 (0) | 1 (1) | 3 (12) |
| No recall | 266 (47) | 67 (52) | 50 (52) | 26 (30) | 20 (43) | 17 (52) | 15 (52) | 13 (45) | 48 (55) | 10 (40) |
| **test com-**<br>**bination**<br>**done<sup>d</sup>** | 232 (36) | 48 (35) | 49 (38) | 53 (59) | 14 (31) | 7 (18) | 16 (44) | 6 (22) | 30 (30) | 9 (28) |

nNO, nasal nitric oxide. HSVA, high-speed video microscopy. EM, electron microscopy. Missing values and the answer category “I don’t know” were classified as “no recall” (missing values: nNO: n = 16, biopsy: n = 6, genetic testing: n = 9, HSVA: n = 1, EM: n = 1). <sup>a</sup>Countries with N≥25 displayed in table, countries with N<25 were categorised into other European countries and other non-European countries. <sup>b</sup>Only participants age ≥ 5 years are included. <sup>c</sup>Proportions of HSVA and EM are calculated out of people who report a biopsy. <sup>d</sup>Tests combination refers to nNO, biopsy and genetics performed, only participants age ≥ 5 years who report any diagnostic test done were included (n = 637).

**S4 Table.** Factors associated with nNO measurement, biopsy and genetic tests, in people with primary ciliary dyskinesia (PCD) (COVID-PCD study).

|  | **nNO<sup>a</sup>**<br>n = 671<br>OR (95%CI) | **Biopsy**<br>n = 724<br>OR (95%CI) | **Genetic testing**<br>n = 724<br>OR (95%CI) |
| --- | --- | --- | --- |
| **Year of diagnosis**<br>(reference category: < 2001) |  |  |  |
| 2001-2010 | 1.5 (0.94-2.3) | 1.95 (1.2-3.2) | 1.6 (1.1-2.6) |
| > 2010 | 2.2 (1.5-3.2) | 3.2 (2.1-4.9) | 4.7 (3.2-6.9) |
| **Situs abnormalities**<br>(reference category: no) |  |  |  |
| yes | 0.5 (0.4-0.7) | 0.5 (0.4-0.8) | 0.7 (0.5-0.94) |
| **Countries/regions**<br>(reference category: United Kingdom) |  |  |  |
| North America | 0.8 (0.5-1.3) | 0.2 (0.1-0.4) | 2.1 (1.3-3.5) |
| Germany | 1.8 (1.03-3.2) | 0.6 (0.3-1.3) | 1.9 (1.1-3.4) |
| Switzerland | 0.5 (0.2-1.1) | 0.4 (0.2-0.8) | 0.6 (0.3-1.2) |
| Italy | 1.005 (0.5-2.0) | 1.4 (0.5-4.1) | 1.3 (0.7-2.7) |
| France | 0.7 (0.3-1.5) | 0.2 (0.1-0.5) | 2.3 (1.1-5.5) |
| Australia | 0.7 (0.3-1.6) | 0.97 (0.3-3.6) | 0.7 (0.3-1.7) |
| Other European countries | 0.7 (0.4-1.4) | 0.4 (0.2-0.8) | 1.4 (0.8-2.4) |
| Other non-European countries | 0.4 (0.2-0.95) | 0.2 (0.1-0.5) | 0.6 (0.3-1.4) |

Abbreviations: nNO, nasal nitric oxide. Odds Ratio (OR) and 95% Confidence Interval (CI) reported. Odds Ratios were adjusted for all variables included in the multivariable model. Performed tests: Participants who report that the test was performed ("yes") were compared to the group who reported either no test ("no") or did not recall the test ("I don't know" and missing). <sup>a</sup>Only participants age ≥ 5 years are included.

**S5 Table.**
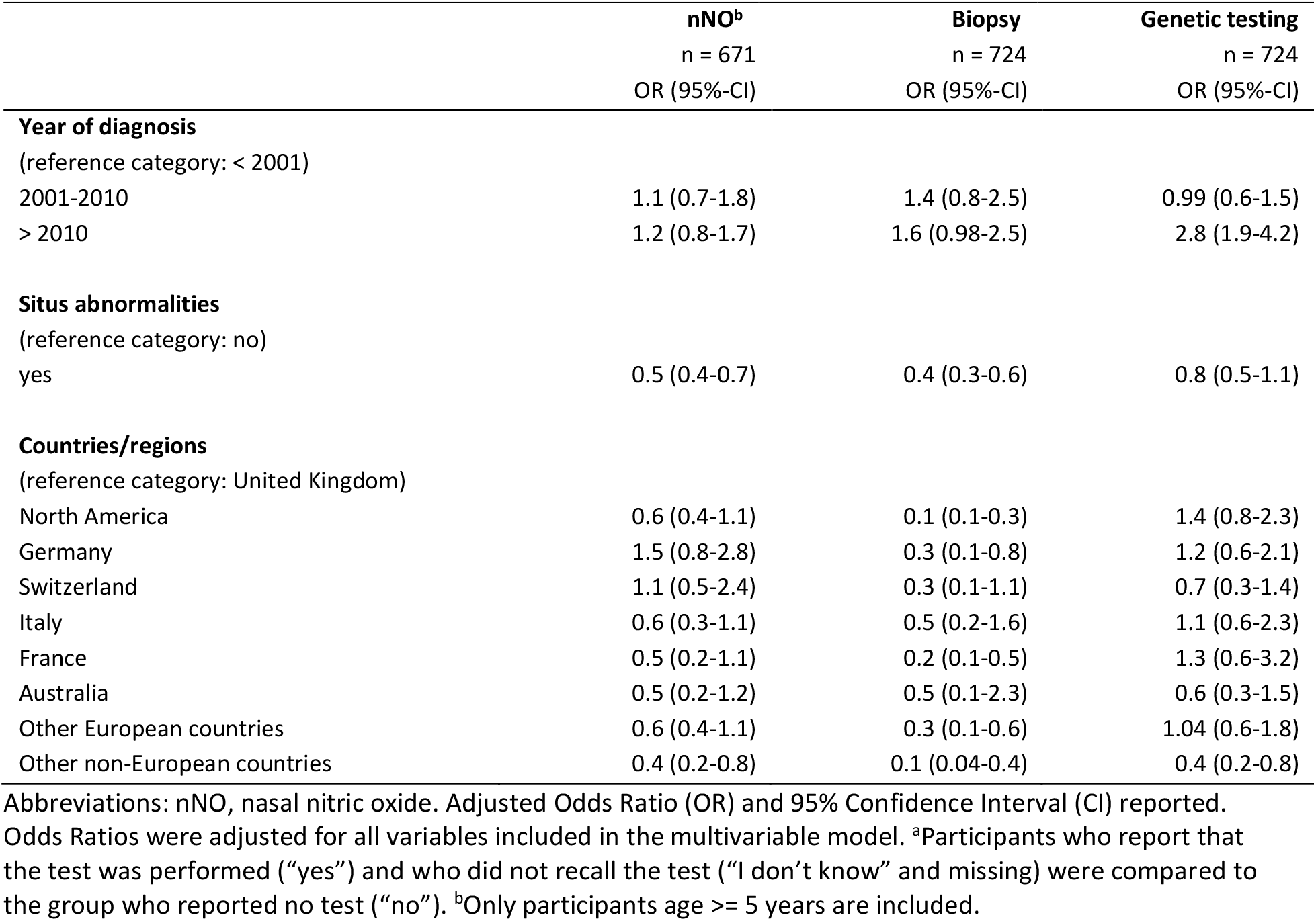
Sensitivity analysis of factors associated with performed nNO, biopsy and genetic testing in which we assumed that everybody in the group “no recall” had the test done in the COVID-PCD studya.

|  | nNO<sup>b</sup><br>n = 671<br>OR (95%-CI) | Biopsy<br>n = 724<br>OR (95%-CI) | Genetic testing<br>n = 724<br>OR (95%-CI) |
| --- | --- | --- | --- |
| **Year of diagnosis**<br>(reference category: < 2001) |  |  |  |
| 2001-2010 | 1.1 (0.7-1.8) | 1.4 (0.8-2.5) | 0.99 (0.6-1.5) |
| > 2010 | 1.2 (0.8-1.7) | 1.6 (0.98-2.5) | 2.8 (1.9-4.2) |
| **Situs abnormalities**<br>(reference category: no) |  |  |  |
| yes | 0.5 (0.4-0.7) | 0.4 (0.3-0.6) | 0.8 (0.5-1.1) |
| **Countries/regions**<br>(reference category: United Kingdom) |  |  |  |
| North America | 0.6 (0.4-1.1) | 0.1 (0.1-0.3) | 1.4 (0.8-2.3) |
| Germany | 1.5 (0.8-2.8) | 0.3 (0.1-0.8) | 1.2 (0.6-2.1) |
| Switzerland | 1.1 (0.5-2.4) | 0.3 (0.1-1.1) | 0.7 (0.3-1.4) |
| Italy | 0.6 (0.3-1.1) | 0.5 (0.2-1.6) | 1.1 (0.6-2.3) |
| France | 0.5 (0.2-1.1) | 0.2 (0.1-0.5) | 1.3 (0.6-3.2) |
| Australia | 0.5 (0.2-1.2) | 0.5 (0.1-2.3) | 0.6 (0.3-1.5) |
| Other European countries | 0.6 (0.4-1.1) | 0.3 (0.1-0.6) | 1.04 (0.6-1.8) |
| Other non-European countries | 0.4 (0.2-0.8) | 0.1 (0.04-0.4) | 0.4 (0.2-0.8) |

Abbreviations: nNO, nasal nitric oxide. Adjusted Odds Ratio (OR) and 95% Confidence Interval (CI) reported.
Odds Ratios were adjusted for all variables included in the multivariable model. <sup>a</sup>Participants who report that the test was performed (“yes”) and who did not recall the test (“I don’t know” and missing) were compared to the group who reported no test (“no”). <sup>b</sup>Only participants age ≥ 5 years are included.

**S6 Table.** Performance of nNO measurement, biopsy and genetic tests by year of diagnosis, situs abnormalities and countries, in people with primary ciliary dyskinesia (PCD) (COVID-PCD study).

|  | nNO done<sup>a</sup><br>n = 342<br>n (%) | No nNO<br>done<sup>a</sup><br>n = 351<br>n (%) | Brushing<br>done<br>n = 561<br>n (%) | No brushing<br>done<br>n = 186<br>n (%) | Genetics<br>done<br>n = 435<br>n (%) | No genetics<br>done<br>n = 312<br>n (%) |
| --- | --- | --- | --- | --- | --- | --- |
| **Year of diagnosis** |  |  |  |  |  |  |
| < 2001 | 85 (37) | 146 (63) | 145 (63) | 86 (37) | 89 (39) | 142 (61) |
| 2001-2010 | 69 (47) | 77 (53) | 111 (76) | 35 (24) | 74 (51) | 72 (49) |
| > 2010 | 176 (60) | 118 (40) | 291 (84) | 56 (16) | 261 (75) | 86 (25) |
| Missing | 12 (58) | 10 (42) | 14 (60) | 9 (40) | 11 (58) | 12 (52) |
| **Situs abnormalities** |  |  |  |  |  |  |
| No | 219 (57) | 164 (43) | 326 (81) | 76 (19) | 262 (65) | 140 (35) |
| Yes | 123 (40) | 187 (60) | 235 (68) | 110 (32) | 173 (71) | 172 (29) |
| **Countries/regions** |  |  |  |  |  |  |
| United Kingdom | 76 (52) | 69 (48) | 128 (85) | 22 (15) | 76 (51) | 74 (49) |
| North America | 68 (48) | 74 (52) | 97 (61) | 61 (39) | 108 (68) | 50 (32) |
| Germany | 66 (66) | 34 (34) | 87 (81) | 20 (19) | 73 (68) | 34 (32) |
| Switzerland | 15 (35) | 28 (65) | 33 (69) | 15 (31) | 18 (38) | 30 (62) |
| Italy | 24 (49) | 25 (51) | 46 (87) | 7 (13) | 28 (53) | 25 (47) |
| France | 19 (46) | 22 (54) | 29 (66) | 15 (34) | 31 (70) | 13 (30) |
| Australia | 13 (46) | 15 (54) | 29 (88) | 4 (12) | 16 (48) | 17 (52) |
| other European countries | 47 (44) | 59 (56) | 87 (76) | 28 (24) | 68 (59) | 47 (41) |
| other countries | 14 (36) | 25 (64) | 25 (64) | 14 (36) | 17 (44) | 22 (56) |

Abbreviations: nNO, nasal nitric oxide. Performed tests: Participants who report that the test was performed ("yes") were compared to the group who reported either no test ("no") or did not recall the test ("I don't know" and missing). <sup>a</sup>Only participants age ≥ 5 years are included.

